# A novel framework leveraging non-causal associations reveals shared pathways linking inflammation and cancer risk

**DOI:** 10.64898/2026.08.30.26361622

**Authors:** James Yarmolinsky, Francesca R Cavallo, Fotios Koskeridis, Xinzhu Yu, Emmanouil Bouras, George Richenberg, Ilaria Costantini, Devleena Ray, Benjamin Woolf, Ville Karhunen, Leila Ellis, Philip C Haycock, Gibran Hemani, George Davey Smith, Kostas K Tsilidis, Verena Zuber, James D McKay, Abbas Dehghan, Ioanna Tzoulaki

**Author notes:** **Corresponding author:** James Yarmolinsky, PhD, Department of Epidemiology and Biostatistics, School of Public Health, Imperial College London, London, UK.

## Abstract

Confounding is a central challenge in observational studies. Here, we propose a framework for identifying confounders of two non-causally related traits by employing cross-trait pleiotropy analysis to detect genetic loci that affect both traits and multi-trait colocalisation to identify molecular phenotypes mediating these effects. We apply this approach to the analysis of C-reactive protein (CRP) - a non-specific marker of inflammation - and 10 inflammation-related cancers. In UK Biobank, higher pre-diagnostic CRP levels are associated with increased risk of multiple cancers, but bidirectional Mendelian randomization provides little evidence for a causal relationship. Cross-trait genetic analyses identify 92 loci with shared CRP-cancer effects including those with established roles in cancer and 50 novel loci such as *RSPO3* (breast cancer) and *GCKR* (colorectal cancer). Integration with proteomic and single-cell transcriptomic data identified putative molecular mediators at 24 loci including plasma TLR1 levels in breast cancer and CD4^+^ T cell *IRF5* expression in kidney cancer. Notably, 15 candidate effector genes encode targets of approved or investigational medications, including *IL6*, *PDE4D*, and *CASP8*, indicating potential opportunities for their repurposing for cancer prevention. The proposed approach provides a generalisable framework for leveraging non-causal phenotypic relationships to yield insights into disease mechanisms and therapeutic targets for disease prevention.

## Introduction

Confounding, a bias due to the presence of common causes of exposures and outcomes, is a central limitation of epidemiological research^1^. Observational studies typically use statistical adjustment to control for confounding factors^2,3^. However, where confounders are unknown, known but unmeasured, or measured with error, epidemiological associations will remain biased^4,5^. Genetic epidemiological approaches that use genetic variants as proxies for environmental and molecular exposures, such as Mendelian randomization (MR), have emerged as an alternative tool to strengthen causal inference in observational settings^6-8^. These approaches leverage the natural randomisation of germline genetic variants at meiosis and conception to minimise confounding and have been increasingly applied to the discovery of modifiable disease risk factors, the elucidation of disease biology, and the prioritisation of novel drug targets^9-13^.

While genetic approaches are typically used to mitigate confounding in observational data, it is underappreciated that genetics can also provide a tool for discovering phenotypic confounders of observational associations. Specifically, where two traits are phenotypically correlated but not causally related, the identification of genetic variants with shared causal effects on both traits can provide insight into the confounders underlying their phenotypic correlation. This is because of the parallel underlying structure of phenotypic confounders and shared causal variants impacting two non-causally related traits as *common causes* of both traits.

C-reactive protein (CRP) is an acute-phase protein widely used as a marker of systemic inflammation^14-16^. Elevated CRP levels have been consistently associated with increased risk of multiple cancers in observational studies^17-26^. However, genetic evidence does not support a causal role of CRP in cancer development, suggesting that observational associations may be confounded by unknown or unmeasured factors upstream to CRP^27-32^. The identification of these confounders could provide insight into inflammation-related mechanisms contributing to cancer risk and potential novel therapeutic targets for cancer prevention.

Here, we propose a framework that leverages cross-trait genetic pleiotropy and multi-trait colocalisation to identify shared molecular mechanisms underlying non-causal phenotypic associations. We first illustrate the equivalence between phenotypic confounders and shared causal variants where two traits are phenotypically correlated but not causally related. Applying this approach to CRP and 10 inflammation-related cancers, we demonstrate that their phenotypic association is unlikely to be causal, and then systematically identify shared genetic variants and, through these, molecular intermediates influencing both traits. Finally, we map candidate effector genes and molecular phenotypes that influence CRP and cancer to approved and investigational medications and explore their translational relevance for cancer prevention. The proposed approach provides a generalisable framework for leveraging non-causal phenotypic relationships to yield novel insights into disease mechanisms and therapeutic targets for disease prevention.

## Results

### Overview of methodological approach

Our approach relies on the assumption of the equivalence between shared causal variants and phenotypic confounders for two traits that are not causally-related. To illustrate their equivalence, consider a hypothetical example where a single causal variant (*X*) influences two traits (*Y*, *Z*).

There are three potential configurations that may explain the relationship between the variant and these traits: i) variant *X* influences trait *Y* that in turn influences trait *Z* (“mediated pleiotropy”), ii) variant *X* influences trait *Z* that in turn influences trait *Y* (“mediated pleiotropy”), and iii) variant *X* influences traits *Y* and *Z* through distinct pathways (“biological pleiotropy”)(**Fig 1A**). In the setting where traits *Y* and *Z* are phenotypically correlated but not causally related, a shared causal variant influencing both traits is only compatible with configuration iii. Assuming gene-environment equivalence (see **Fig 1** footnote), the identification of a shared causal variant for *Y* and *Z* combined with an understanding of the molecular mechanism through which it operates can provide insight into shared phenotypic mechanisms influencing both traits (**Fig 1B**)^33^. Hence, when two traits are phenotypically correlated but not causally related to each other, we can leverage the genetic basis of their phenotypic association to learn about confounders underlying this association.

**Figure 1.**
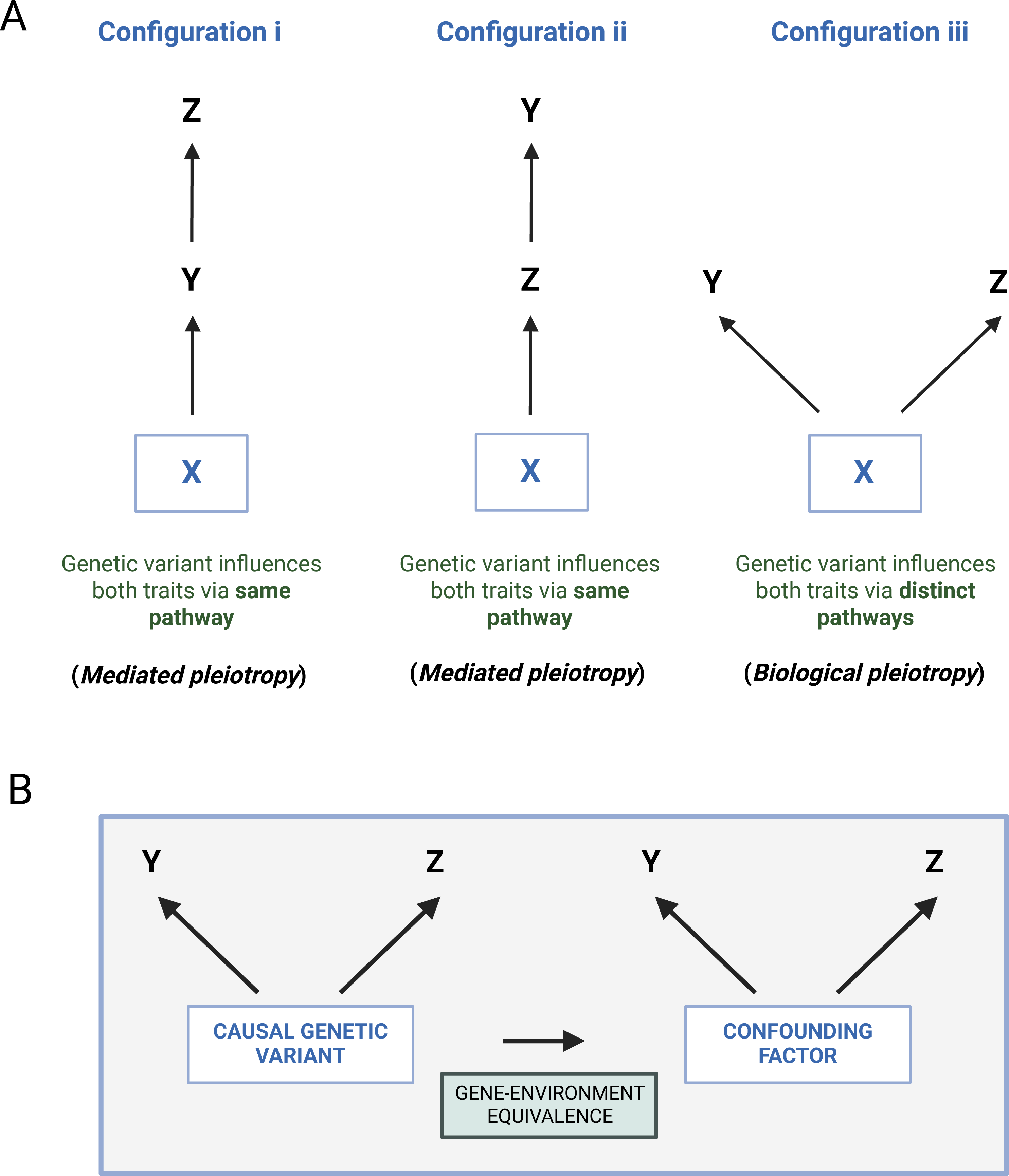
Configurations through which a genetic variant can causally influence two traits. Gene-environment equivalence assumes that the perturbation of a phenotype through genetic or environmental means will confer the same effect on a downstream trait

For example, cigarette smoking is a common cause of coffee intake and coronary artery disease, generating a non-causal association between these traits (**Fig 2A**)^34-37^. A single-nucleotide polymorphism (SNP, rs1051730) within the *CHRNA5-A3-B4* gene cluster has been shown to increase smoking heaviness, coffee intake, and atherosclerosis risk in carriers of the A allele (**Fig 2B**)^38^. The location of this variant within a gene cluster strongly implicated in nicotine dependence can be used to infer that it likely has a primary effect on smoking behaviour with subsequent downstream effects on coffee intake and atherosclerosis risk. Clinical trial and genetic evidence do not support an atherogenic effect of coffee intake and a higher genetic liability to atherosclerosis increasing subsequent coffee intake is implausible, suggesting that mediated pleiotropy (configurations i or ii) is unlikely^39-42^. The most likely explanation for the shared effect of rs1051730 on these three traits is that genetically proxied cigarette smoking influences coffee intake and atherosclerosis risk through distinct pathways (configuration iii) and that, phenotypically, cigarette smoking is a confounder of both traits.

**Figure 2.**
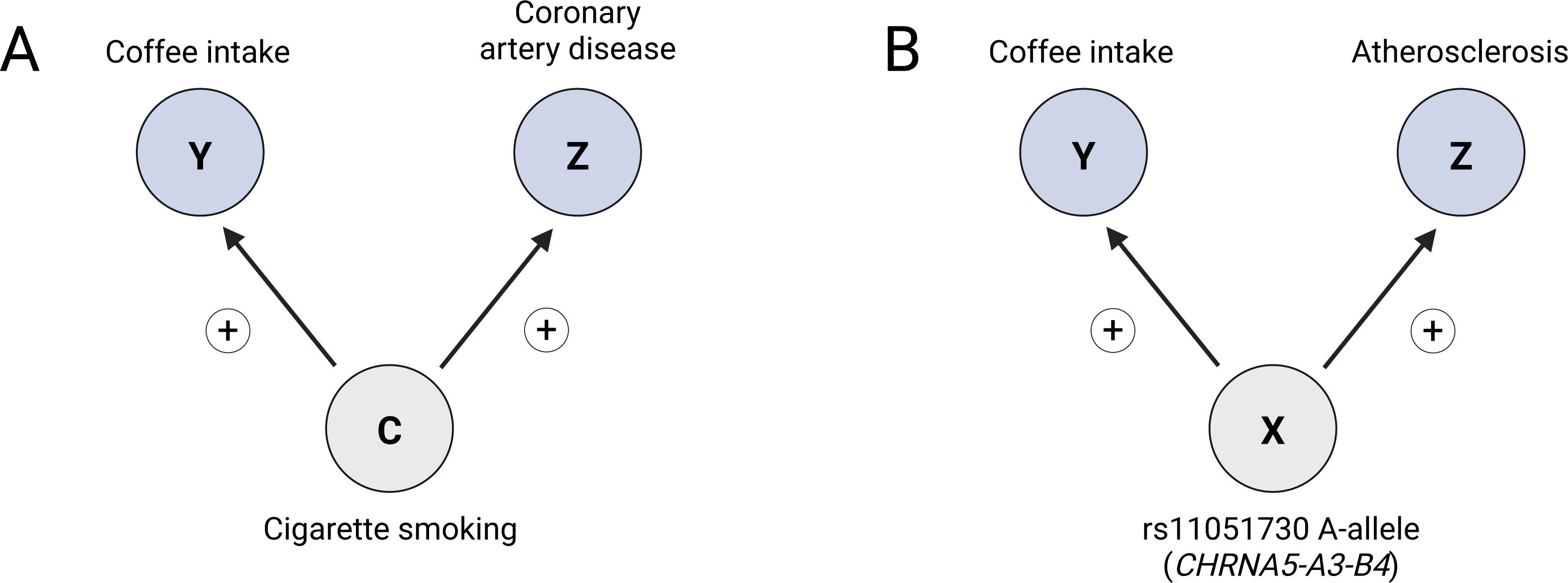
Demonstration of equivalence of phenotypic confounders and shared causal variants using cigarette smoking, coffee intake, and cardiovascular endpoints as an exemplar.

We hypothesized that genetic evidence could be used to recapitulate confounding structures underlying the phenotypic association between CRP and cancer and provide insight into inflammation-related mechanisms influencing cancer development. To test this, we first establish the absence of a causal relationship between CRP and cancer risk using bidirectional Mendelian randomization. We then apply cross-trait pleiotropy analysis and genetic colocalisation to identify shared genetic signals of CRP and cancer risk, followed by integration of proteomic and transcriptomic data to prioritise candidate molecular mediators. A schematic overview of the analytical framework is shown in **Fig 3**.

**Figure 3.**
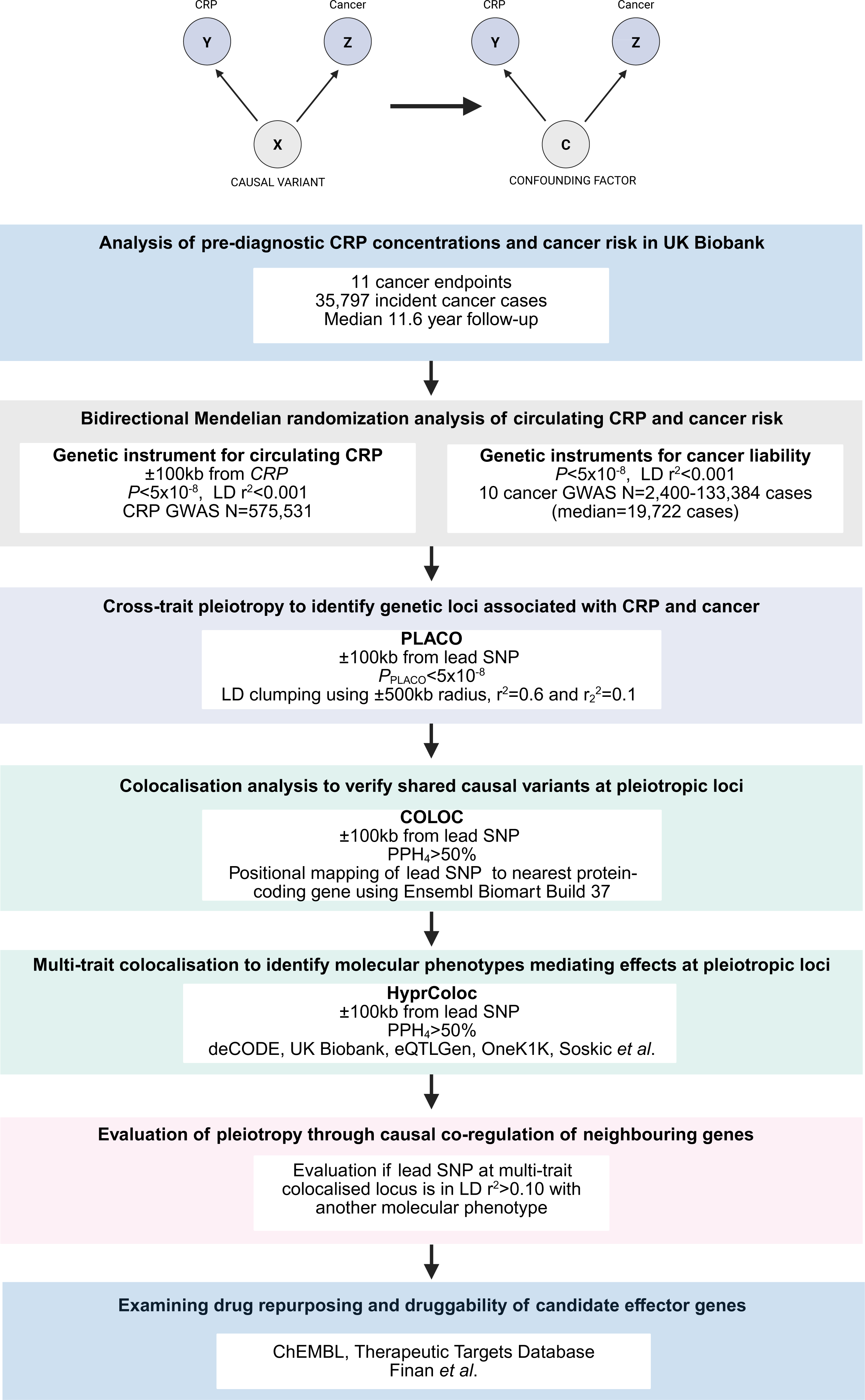
Schematic overview of the analytical framework of this study. GWAS = genome-wide association study, LD = linkage disequilibrium

### Analysis of pre-diagnostic CRP concentrations and cancer risk in UK Biobank

To verify the phenotypic correlation between CRP and cancer risk, we first examined the prospective association of pre-diagnostic CRP concentrations with risk of 11 cancers previously linked to chronic inflammation in the UK Biobank (**Table 1**)^43^.

**Table 1.** Characteristics of UK Biobank prospective cohort study participants by quartiles of circulating C-reactive protein (CRP) concentrations (N=362,638)

| Characteristic | C-reactive protein concentrations |  |  |  |
| --- | --- | --- | --- | --- |
|  | Q1<br>(N=91629) | Q2<br>(N=90004) | Q3<br>(N=90589) | Q4<br>(N=90416) |
| Females (%) | 50193 (54.8) | 45880 (51.0) | 47498 (52.4) | 53469 (59.1) |
| Age, mean (SD) | 71.1 (13.2) | 76.4 (14.3) | 80.1 (15.3) | 84.5 (17.5) |
| Body mass index, mean (SD) | 24.7 (3.4) | 26.6 (3.7) | 28.1 (4.2) | 30.2 (5.6) |
| Height, mean (SD) | 169.4 (9.2) | 169.2 (9.3) | 168.5 (9.3) | 167.1 (9.2) |
| Deprivation index, mean (SD) | -1.5 (2.9) | -1.5 (3.0) | -1.4 (3.0) | -1.0 (3.2) |
| Max days active, mean (SD) | 5.7 (1.7) | 5.6 (1.7) | 5.6 (1.8) | 5.4 (1.9) |
| White ethnicity, n (%) | 86627 (94.5) | 85360 (94.8) | 85880 (94.8) | 85086 (94.1) |
| Ever smoked, n (%) | 35895 (39.2) | 38348 (42.6) | 41072 (45.3) | 44629 (49.4) |
| Has alcohol weekly, n (%) | 68193 (74.4) | 65253 (72.5) | 62668 (69.2) | 57021 (63.1) |
| Family history of cancer, n (%) | 27941 (30.5) | 27756 (30.8) | 28700 (31.7) | 28497 (31.5) |

Quartiles are per natural log-transformed standardised baseline CRP levels. Deprivation Index = Townsend Deprivation Index.

Over a median follow-up of 11.6 years (35,797 incident cases), higher CRP levels were linked to increased risk of 10 of 11 inflammation-related cancers (**Fig 4A**), with no evidence of association for ovarian cancer. These associations remained consistent after adjustment for socioeconomic, anthropometric, and lifestyle factors, and after excluding the first three years of follow-up. The 10 cancers observationally linked to CRP were taken forward to downstream genetic analyses.

**Figure 4.**
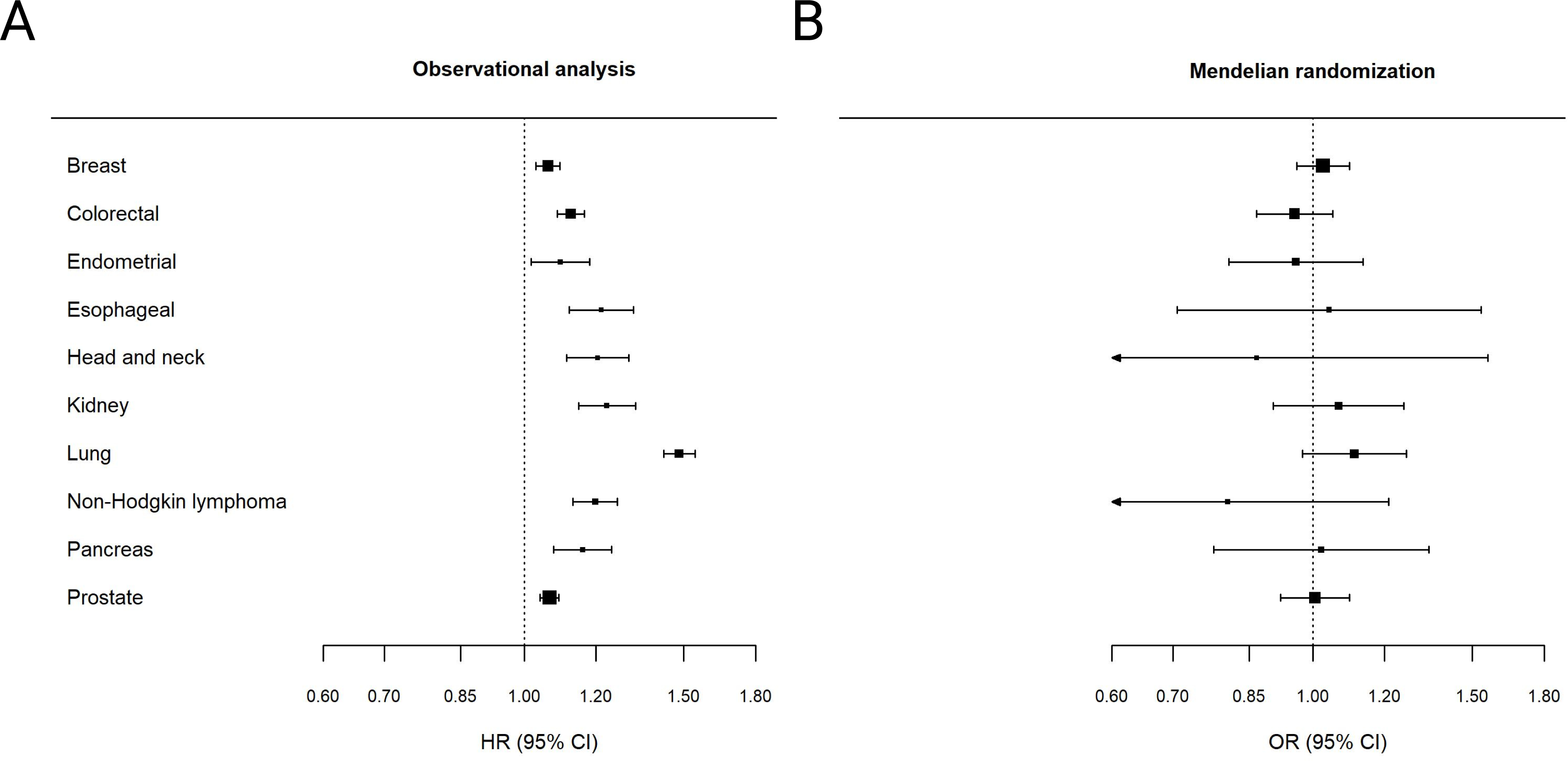
Forest plots comparing phenotypic association of pre-diagnostic directly measured CRP and cancer risk with cis-Mendelian randomization estimate of the effect of circulating CRP on cancer risk. HR = hazard ratio, SNP = single-nucleotide polymorphism, OR = odds ratio

### Evaluating causal relationships between CRP and cancer risk using bidirectional Mendelian randomization analysis

To evaluate if there is evidence to support a causal relationship between circulating CRP and cancer risk, we then performed bidirectional Mendelian randomization (MR) analysis using inverse-variance weighted (IVW) models and pleiotropy-robust methods in sensitivity analyses (see **Methods**). F-statistics for genetic instruments for CRP (F=7,924) and genetic liability to cancer (F=30-41,547) suggested that findings were unlikely to be influenced by weak instrument bias (**Table S1**).

In *cis*-MR evaluating the effect of circulating CRP on cancer risk, we found no evidence to support a causal effect of CRP across 10 cancer sites (**Figure 4B**, **Table S2**). In reverse MR analyses examining the effect of genetic liability to cancer on CRP levels, we did not detect an association for 9 of 10 cancers evaluated (**Table S3**). In IVW models there was evidence for an effect of genetic liability to lung cancer on CRP (β=0.05, 95% CI:0.02 to 0.07, *P*=6.48x10^-4^). However, in models accounting for correlated pleiotropy these associations attenuated toward the null (CAUSE: β=0.01, 95% credible interval:0.00 to 0.03, ΔELPD_SharingvsCausal_=-0.51, *P*=0.36; MR-CUE: β=0.01, 95% CI:-0.01 to 0.03, *P*=0.25)(**Table S4**). In addition, 8 of 13 lung cancer susceptibility SNPs with data on lifetime smoking index were associated with this index (*P*<0.05) and adjustment for smoking using multivariable MR likewise generated little evidence of association of lung cancer liability with CRP (β=-0.12, 95% CI:- 0.39 to 0.15, *P*=0.38). These findings suggest that genetic liability to lung cancer is unlikely to causally impact CRP levels and that IVW estimates are potentially biased by correlated pleiotropy via smoking behaviour.

### Cross-trait pleiotropy analysis to identify genetic loci associated with CRP and cancer

Having established that CRP and risk of 10 cancers are phenotypically correlated but unlikely to be causally related, we then performed cross-trait pleiotropy analysis to identify genetic variants associated with CRP and 10 cancers using pleiotropic analysis under a composite null hypothesis (PLACO)(**Methods**)^44^. In pair-wise analyses, we identified 32,567 SNPs associated with circulating CRP and cancer (*P*_PLACO_<5x10^-8^). After clumping these SNPs, there were 458 distinct loci (**Figure 5**, **Tables S5-14**).

**Figure 5.**
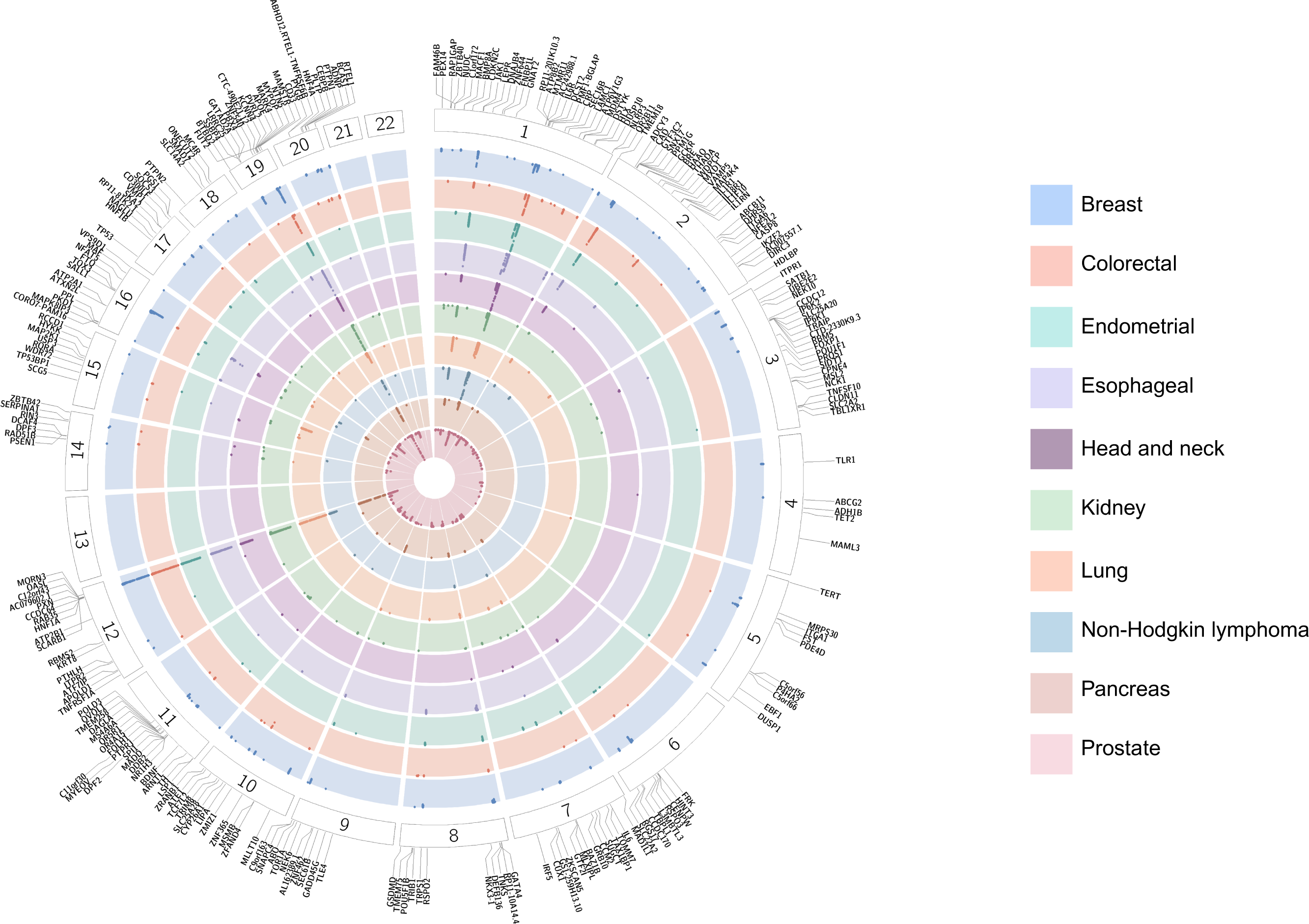
Circular plot with pleiotropic analysis under composite null hypothesis (PLACO) results for analyses examining pleiotropic loci for CRP and cancer. Each inner track is a Manhattan plot of CRP-cancer associations (*P*_PLACO_<5x10^-8^) presented per cancer site. Labelled genes represent the closest gene to the lead SNP at each independent pleiotropic locus.

### Genetic colocalisation to verify shared causal variants underlying CRP-cancer associations

Genetic associations at these 458 loci are compatible with CRP and cancer being influenced by i) the same underlying causal variant and ii) distinct causal variants that are in linkage disequilibrium (LD). To distinguish between these scenarios, we performed pairwise colocalisation analyses of CRP and cancer associations on the genomic region (±100 kb) centred on the lead SNP at each locus using the coloc package (**Methods**). We identified 92 loci with evidence of a single shared causal variant influencing CRP and cancer (PPH_4_> 50%)(**Table S15**). Cancers of the breast (23), prostate (20), and kidney (13) had the most pleiotropic loci with colocalisation evidence. 9 loci were shared across two or more cancer outcomes, including those mapped to *CENPW* (head and neck, pancreatic), *TMEM18* (breast, prostate), and *SERPINA1* (non-Hodgkin lymphoma, prostate).

### Recapitulation of known risk factor-cancer incidence relationships

Among these loci we identified several proof-of-principle examples supporting the validity of our approach. For example, we identified a shared genetic signal between CRP and lung cancer at the *CHRNA5* locus (PPH_4_=88.7%). *CHRNA5* encodes the nicotinic acetylcholine receptor α5 subunit which is implicated in nicotine dependence, in turn impacting smoking behaviour, a driver of low-grade inflammation and established risk factor for lung cancer^45-47^. The most likely causal variant at this locus (rs55781567, 78.8% PP explained by SNP) is a SNP in the 5’-UTR of the *CHRNA5* gene previously reported to increase cigarette smoking frequency and duration in carriers of the G allele (**Fig 6A**)^38,48^. We found that each copy of the G allele of this variant also raised CRP levels (β=0.01, *P*=2.10x10^-7^) and increased lung cancer risk (OR=1.30, *P*=3.08x10^-103^). Our genetic findings recapitulating the established effects of smoking on CRP and lung cancer risk thus support cigarette smoking as a confounder of their phenotypic relationship.

**Figure 6.**
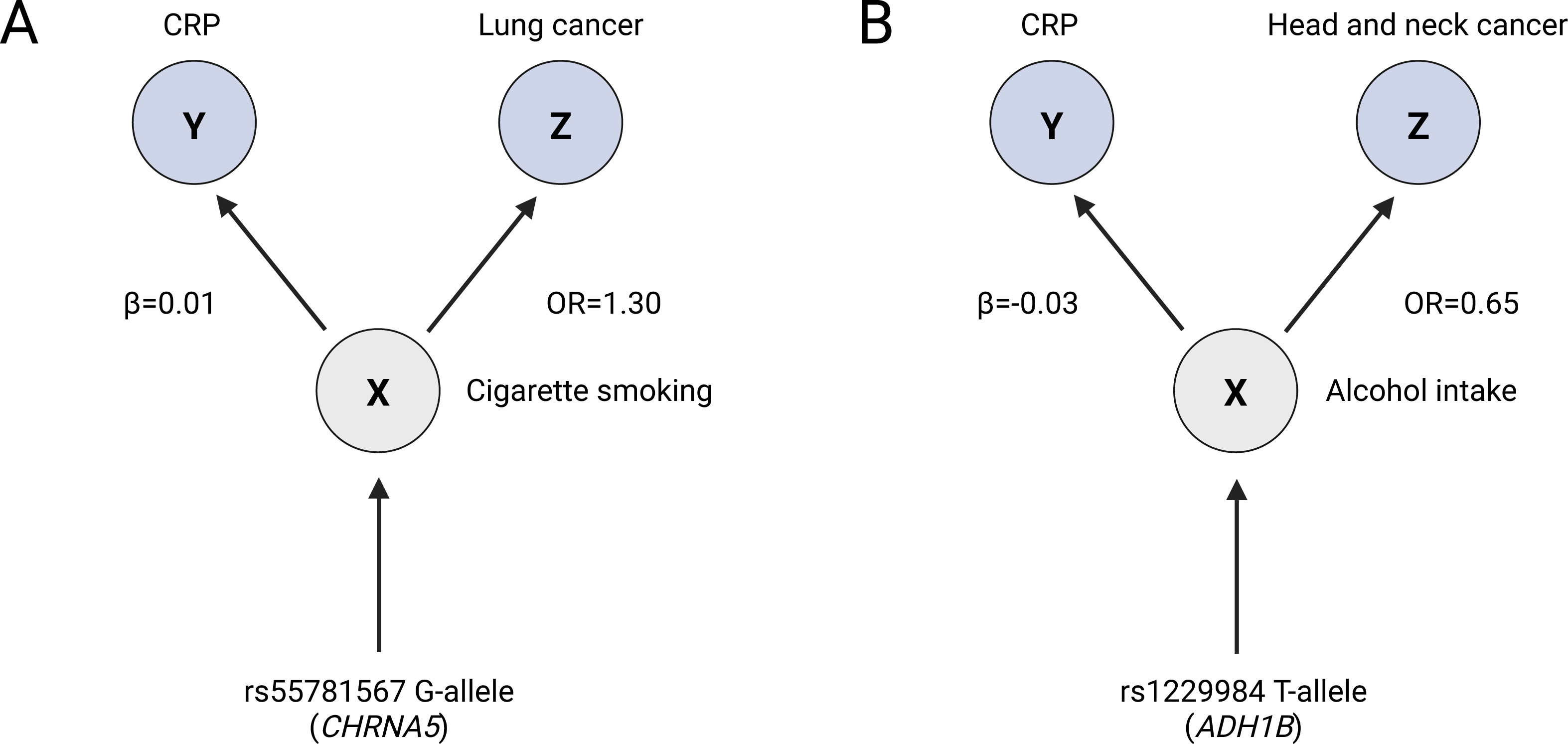
Examples of recapitulation of known risk factor-cancer onset relationships among pleiotropic colocalised loci influencing CRP and cancer.

We also found a shared genetic signal between CRP and head and neck cancer at the *ADH1B* locus (PPH_4_=55.7%). *ADH1B* encodes the alcohol dehydrogenase 1B enzyme which plays a critical role in metabolism and subsequent consumption of alcohol, a contributor to chronic inflammation and key risk factor for head and neck cancer^49,50^. The most likely causal variant at this locus (rs1229984, 99.9% PP explained by SNP) is a missense variant previously shown to lower levels of alcohol consumption in carriers of the T allele^51^. In our analyses we found that each copy of the T allele of this variant also lowered CRP levels (β=-0.03, *P*=7.90x10^-6^) and reduced head and neck cancer risk (OR=0.65, *P*=4.28x10^-14^)(**Fig 6B**). Our findings indicating a shared genetic effect of this *ADH1B* variant on CRP levels and head and neck cancer risk align with the known effect of alcohol consumption on both traits and suggest alcohol as a confounder of their phenotypic relationship.

### Identification of novel cancer susceptibility loci

Along with recapitulating known relationships between established risk factors and cancer onset, we identified 50 novel cancer susceptibility loci across 10 anatomical sites. For example, we found a shared genetic signal between CRP and breast cancer at the *RSPO3* locus (PPH_4_=93.0%), likely driven by the intronic variant rs72959041 (97.4% PP explained by SNP). G-allele carriers at rs72959041 had both higher CRP levels (β =0.05, *P*=1.94x10^-26^) and an increased breast cancer risk (OR=1.06, *P*=9.44x10^-5^). RSPO3 encodes R-spondin-3, a secreted protein and amplifier of canonical Wnt/β-catenin signaling that has been reported to drive mammary adenocarcinomas in mouse models^52,53^.

We also found a shared signal between CRP and esophageal cancer at the *LRRC25* locus (PPH_4_=96.3%). The most likely causal variant is a SNP 5,162 base pairs (bp) upstream from the *LRRC25* gene (rs1985157, PP SNP=98.0%). Carriers of the T allele had lower CRP levels (β=-0.017, *P*=7.97x10^-18^) and an increased risk of esophageal cancer (OR=1.12, *P*=7.01x10^-5^). Deficiency or knockdown of *LRRC25*, which encodes a critical negative regulator of innate immunity, has been shown to promote anti-tumour immunity and to suppress tumour growth in murine cancer models^54,55^.

Likewise, we identified a shared signal between CRP and pancreatic cancer at the *HNF1A* locus (PPH_4_=92.8%). The most likely causal variant (rs7979478) is an intronic variant in *HNF1A* that reduced CRP (β=-0.1497, *P*<5x10^-324^) and increased pancreatic cancer risk (OR=1.10, *P*=5.89 x 10^-6^) in A-allele carriers. HNF1A is a transcription factor that is highly expressed in pancreatic beta cells and has been shown to drive tumourigenesis and regulate pancreatic cancer stem cell properties in models of pancreatic cancer^56^.

### Multi-trait colocalisation identifies molecular phenotypes mediating effects at pleiotropic loci

CRP is produced by hepatocytes in the liver and subsequently secreted into the bloodstream in response to circulating pro-inflammatory cytokines and other inflammatory signals. To identify upstream molecular phenotypes driving effects on plasma CRP expression and cancer risk at each of the 90 colocalised loci, we next performed multi-trait colocalisation integrating whole blood gene expression and plasma protein expression data for genes neighbouring each locus.

We detected evidence for a shared causal variant influencing a molecular phenotype, CRP, and cancer risk at 27 loci (**Table S16**). At 7 loci, there was multi-trait colocalisation with two or more molecular phenotypes, thus preventing us from confidently assigning a putative molecular intermediate. However, for the remaining 20 loci we were able to infer the potential molecular phenotype mediating the shared genetic signal between CRP and cancer. Of these 20 loci, 13 were identified for prostate, breast, or colorectal cancer; 2 each were identified for head and neck, pancreatic, and endometrial cancer; and 1 was identified for kidney cancer.

For example, we identified a shared genetic signal between plasma TLR1 expression levels, CRP, and breast cancer at the *TLR1* locus (PPH_4_=89.1%), implicating TLR1 as an upstream driver of CRP and breast cancer (**Figure 7**). The most likely causal variant at this locus (rs5743618, 100% PP explained by SNP) increased plasma TLR1 levels (β=0.138, *P*=2.41x10^-61^), CRP levels (β=0.015, *P*=1.18 x 10^-9^) and breast cancer risk (OR=1.05, *P*=1.13x10^-12^) per copy of the A allele. TLR1 is an innate immune receptor highly expressed in whole blood that forms a heterodimer with TLR2 to detect pathogen-associated molecular patterns and initiate an inflammatory response^57^.

**Figure 7.**
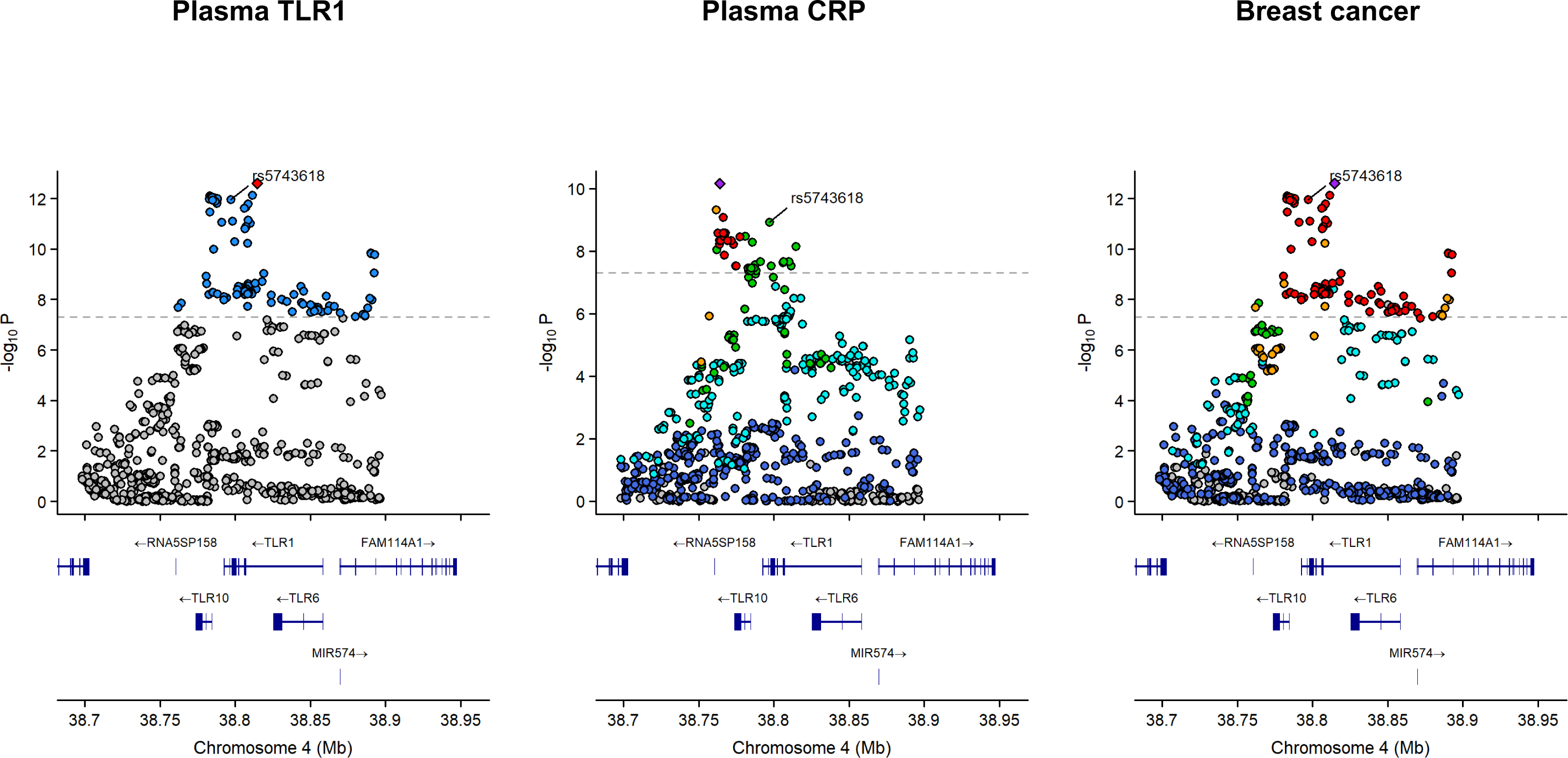
Regional plots showing association of genetic variants with plasma TLR1, plasma CRP, and breast cancer risk in the *TLR1* locus. The putative causal variant underlying multi-trait colocalization at the TLR1 locus (rs5743618) is labelled on each plot.

We found a shared genetic signal between plasma GCKR (alias GKRP), CRP, and colorectal cancer at the *GCKR* locus (PPH_4_=97.3%). The most likely causal variant underlying this association (rs4665972, 100% PP explained by SNP) increased plasma GCKR levels (β=0.08, *P*=5.51x10^-24^) and colorectal cancer risk (OR=1.04, *P*=9.98x10^-6^) but reduced CRP levels (β=-0.07, *P*=9.73x10^-273^) per copy of the C allele. GCKR is an endogenous inhibitor of GCK, the target of the type 2 diabetes medication dorzagliatin and a key regulator of blood glucose homeostasis that has been shown to influence hepatic expression of cytokines upstream to CRP (e.g. interleukin-1β, interleukin-6)^58-60^.

We also found evidence for shared genetic signals influencing plasma SERPINA5 levels, CRP, and prostate and head and neck cancers at *SERPINA5* (PPH_4_=99.9% prostate cancer, 95.9% head and neck cancer). The most likely causal variant at this locus for all four traits (rs28929474, 99.9% PP explained by SNP in both sets of multi-trait colocalisation analyses) increased plasma SERPINA5 levels (β=0.20, *P*=2.16x10^-19^), lowered CRP levels (β=-0.10, *P*=7.50x10^-42^), and reduced risk of both prostate cancer (OR=0.88, *P*=1.67x10^-10^), and head and neck cancer (OR=0.75, *P*=2.73x10^-5^) per copy of the T allele. SERPINA5 is a serine protease inhibitor involved in blood coagulation that has also been shown to inhibit tumour growth by regulating signalling pathways like PI3K/AKT/mTOR^61,62^.

### Single-cell eQTL data reveals immune cell-specific mechanisms influencing cancer risk

Bulk whole blood gene expression data cannot identify if regulatory effects on CRP and cancer are driven by specific immune cell types and may fail to detect evidence of colocalisation of cell type-specific effects. To resolve this issue, we used single-cell data on 14 immune cells (plasma cells, B-cell subsets, CD4^+^ and CD8^+^ T cell subsets, natural killer cells, monocyte subsets, dendritic cells) to identify potential immune cell-type specific mechanisms driving effects on cancer risk. We also used single-cell data on memory and naive CD4^+^ T cell response to activation at 3 time points (16h, 40h, 5 days) to examine potential cell-state specific mechanisms influencing cancer.

We identified 9 loci with evidence of regulatory effects on CRP and cancer being driven by immune cell-type or cell-state-specific mechanisms, of which 7 could be confidently assigned to one molecular phenotype (**Table S17**). For example, single-cell data helped to clarify that a shared genetic signal across whole blood *CENPW* expression, CRP and pancreatic cancer at the *CENPW* locus was likely driven by 16h-stimulated CD4^+^ memory (PPH_4_=57.2%) and CD8^+^ T cell (PPH4=54.1%) *CENPW* expression. CD4 memory T cells play a critical role in the adaptive immune response by providing rapid, enhanced protection against previously encountered pathogens while CD8^+^ T cells target virus-infected, tumour, and abnormal cells^63^.

We also identified blood immune cell-specific effects at 4 loci that were not detected in analyses using bulk expression data, highlighting the enhanced resolution provided by single-cell data. For example, we discovered that the previously identified shared genetic signals across CRP and kidney cancer at *IRF5* in colocalisation analysis was potentially mediated by an effect of peripheral blood 16h-stimulated memory CD4^+^ T cell *IRF5* expression (PPH_4_=81.1%). The most likely causal variant at this locus (rs3778754, 63.4% PP explained by SNP) increased memory CD4^+^ T cell *IRF5* expression (β=0.64, *P*=2.89x10^-8^) and CRP levels (β=0.01, *P*=2.87x10^-8^) but reduced kidney cancer risk (OR=0.92 *P*=1.49x10^-15^) per copy of the G allele. IRF5 is a transcription factor that regulates expression of pro-inflammatory cytokines (e.g. IL-6, TNF) upstream of CRP production and a putative tumour suppressor for several cancers^50,64,65^.

Likewise, we found evidence that the previously identified shared genetic signal across CRP and esophageal cancer at *LRRC25* in colocalisation analysis was potentially mediated by an effect of peripheral blood immature and naïve B cell (B_IN_) *LRRC25* expression (PPH_4_=91.9%). We also found a shared genetic effect on CRP and breast cancer at *L3MBTL3* that was mediated by CD4^+^ T cell expression of *L3MBTL3* (PPH_4_=77.2%). L3MBLTL3 plays a critical role in haematopoietic development and has been linked to metastasis and poor prognosis in breast cancer patients^66^.

### Shared genetic signals highlight drug repurposing opportunities and druggable targets

To identify approved or investigational medications that target candidate effector genes and molecular intermediates we cross-referenced our results with ChEMBL and Therapeutic Target Database. In total, 15 prioritised genes or molecular intermediates are targets of one or more approved or investigational medications, encompassing a wide variety of indications (autoimmune conditions, cancer, cardiovascular disease)(**Table S18**). This included PDE4D (breast cancer), a target of approved PDE4 inhibitors such as apremilast used to treat plaque psoriasis and psoriatic arthritis and in Behçet’s disease^67,68^. MAP2K1 (kidney cancer) encodes the target of MEK inhibitors (e.g. cobimetinib) that are approved to treat malignant melanoma with BRAF^V600^ mutations^69^. In total, 5 targets were implicated in breast cancer (PDE4D, RSPO3, CASP8, OPRL1, ABCG2), 4 for prostate cancer (MC4R, SERPINA5, FST, CYP26A1), 2 for esophageal cancer (ORA1I, RPS6KB2), 2 for lung cancer (IL6, IKZF2), and 1 each for non-Hodgkin lymphoma (JAK1), kidney cancer (MAP2K1), and head and neck cancer (SERPINA5). We also queried the Druggable Genome database and identified a further 9 candidate effector genes or genes that encode candidate molecular phenotypes that are considered druggable (**Table S19**).

## Discussion

Here we propose a novel framework that leverages the genetic basis of non-causal phenotypic associations to gain insight into shared phenotypic mechanisms that confound these associations. Through application to the analysis of CRP and 10 inflammation-related cancers we demonstrate how this framework can provide insight into inflammation-related mechanisms contributing to cancer development, thus helping to prioritise novel therapeutic targets and repurposing opportunities for cancer prevention.

Among the shared genetic signals influencing CRP and cancer that we identified, we discovered several loci where the most likely causal variant resided in or adjacent to a gene that encoded an established risk factor for that cancer. This included a shared genetic signal for CRP and lung cancer in the *CHRNA5* locus that was likely driven by a variant previously implicated in cigarette smoking and a shared signal for CRP and head and neck cancer in *ADH1B* that was likely mediated by a variant known to influence alcohol intake. These examples provide proof-of-principle that an agnostic cross-trait pleiotropy and colocalisation framework can “re-discover” known causes of cancer and CRP and thus indicate plausible phenotypic confounders of the two.

Along with recapitulating known risk factor-cancer relationships, we identified 50 novel cancer susceptibility loci implicating an extensive catalogue of inflammatory-related mediators in cancer risk such as RSPO3 in breast cancer and LRRC25 in esophageal cancer. These findings validate and extend insights from preclinical studies that pharmacological inhibition of RSPO3 inhibits breast cancer cell growth and that *LRRC25* knockdown suppresses tumour growth in murine cancer models, supporting their further prioritisation as cancer prevention targets^52,53,55^. We also identified upstream molecular drivers of CRP and cancer that are not immune regulators but likely influence inflammation through indirect mechanisms. This included evidence implicating molecular phenotypes involved in coagulation (SERPINA1), glucose metabolism (GCKR), LDL cholesterol metabolism (SORT1), and metabolic function SLC2A12), helping to generate novel hypotheses into cancer-related pathways and guide future work.

In multi-trait colocalisation analysis integrating whole blood gene expression and plasma protein expression data, we identified putative molecular intermediates driving effects on CRP and cancer at 20 loci including those with established roles in inflammatory signalling pathways upstream to CRP. For example, we found evidence that breast cancer was influenced by plasma levels of the toll-like receptor TLR1, an innate immune receptor that participates in NF-kB and MAPK cascades that promote cytokine production, in turn stimulating hepatic CRP production. Toll-like receptors have been shown to contribute to the tumour microenvironment in breast cancer and its pathogenesis and knockdown of *TLR1* has been reported to inhibit breast cancer cells, thus providing orthogonal support to our genetic findings for a role of TLR1 in breast cancer risk^70,71^.

To further resolve immune cell-type and cell-state specific mechanisms underlying cancer risk, we integrated peripheral blood single-cell gene expression data on 14 cell types and 3 states, identifying cell-specific regulatory mechanisms driving effects of 7 molecular phenotypes. This included evidence that 16h-stimulated memory CD4^+^ T cell *IRF5* expression influenced kidney cancer and that immature and naïve B cell *LRRC25* expression influenced esophageal cancer. IRF5 and LRRC25 are both highly expressed in immune cells and implicated in innate immunity albeit with opposing roles in activating (IRF5) and suppressing (LRRC25) the immune response^72,73^. IRF5 has previously been reported to act as both a tumour suppressor and proto-oncogene, highlighting a possible complex and context-specific role in cancer development and necessitating further investigation to elucidate its relationship with kidney cancer risk^74^.

15 of the candidate effector genes or molecular intermediates that we identified are targets of approved or investigational medications and an additional 9 are considered “druggable”. This included targets of approved medications used to treat cancer (e.g. MAP2K1, trametinib) and autoimmune conditions (e.g. PDE4D, apremilast) along with investigational medications for coronary artery disease (e.g. IL6, ziltivekimab), chronic liver disease (e.g. CASP8, emricasan), and autoimmune disease (e.g. CYP26A1, talarozole)^75-79^. Though itself not the target of an approved drug, a putative role of plasma GCKR in colorectal cancer risk supports the repurposing potential of the GCK activator dorzagliatin for colorectal cancer prevention, warranting further investigation^60^. Notably, we find genetic evidence supporting the possible role of interleukin-6 in lung cancer development which, to the best of our knowledge, has not been reported previously though we did not find further evidence of interleukin-6 molecular mediation in multi-trait colocalisation analysis. Interleukin-6 is activated by interleukin-1β in the innate inflammatory cytokine cascade and therefore our findings are consistent with the lung cancer protective effect of the interleukin-1β inhibitor canakinumab observed in the CANTOS trial^80^. Recently, the Phase III ZEUS trial failed to identify an effect of the IL-6 monoclonal antibody ziltivekimab on major adverse cardiovascular events in patients with atherosclerotic cardiovascular disease, chronic kidney disease, and inflammation^81^. Ongoing Phase III trials testing ziltivekimab (ARTEMIS, HERMES) and the anti-interleukin-6 monoclonal antibody clazakizumab (POSIBIL6ESKD) for cardiovascular outcomes in different patient populations to the ZEUS trial will provide an additional opportunity to explore the cardioprotective effects of IL-6 inhibition, in turn supporting the further evaluation of their chemopreventive potential for lung cancer^77,82^.

Confounding remains a central concern in observational studies. Along with identifying potential causal mechanisms influencing disease risk our approach also enables the improved design of conventional observational studies by indicating phenotypic confounders that could be incorporated into confounder adjustment strategies. Similar approaches to ours have been proposed but differ in important ways^83-85^. Warwick *et al*. use the non-causal effect of CRP on various health outcomes to identify potential causal roles of genes mapped to SNPs with genome-wide significant *trans*-acting CRP-effects on these outcomes using Mendelian randomization^84^. Likewise, Hamilton *et al*. used bidirectional Mendelian randomization to demonstrate that the phenotypic, but non-causal, association between lower levels of small high-density lipoprotein cholesterol (HDL) particles and increased risk of sepsis was likely driven by confounding by increased interleukin-6 signalling^85^. In contrast to Warwick and Hamilton *et al*., our approach is not an extension to Mendelian randomization but instead uses an agnostic cross-trait pleiotropy and colocalisation framework to detect shared genetic mechanisms influencing traits that are not causally related, and thus potential confounders of their phenotypic association. Similarly, in contrast to our aim to use genetics to identify phenotypic confounders, Sanderson *et al*. have proposed using Steiger filtering to identify heritable confounders in Mendelian randomization studies and then pre-estimation filtering or multivariable MR to generate causal estimates that are unbiased by these confounders^83^. Common complex diseases are typically highly polygenic and therefore the variant-level genome-wide pleiotropy approach proposed here is more pertinent to the discovery of proteomic and transcriptomic causes of disease that may be more strongly shaped by local *cis*-acting mechanisms^86^.

Strengths of our study include proposing a framework that leverages pleiotropic variants influencing non-causally related traits to identify potential confounders underlying their phenotypic association. We employed a robust analytical strategy to systematically identify shared causal variants impacting CRP and cancer and biologically-informed multi-trait colocalisation analyses to identify single-cell and bulk transcriptomic and proteomic mediators driving these effects while attempting to minimise horizontal pleiotropy through causal co-regulation of neighbouring genes. There are several limitations to this analysis. First, the colocalisation analyses that were performed using coloc assumed a single causal variant at each locus and therefore may not have identified instances of colocalisation across conditionally independent causal variants influencing CRP and cancer. We used this approach to permit parallel downstream multi-trait colocalisation analysis, for which all currently available summary data methods make this assumption. Second, though all putative molecular intermediates had supporting multi-trait colocalisation evidence, we cannot rule out the possibility that their effects on CRP and cancer were mediated through cell types or tissues that were not measured. Likewise, where we did not find evidence of multi-trait colocalisation evidence for a particular molecular intermediate, this does not exclude the possibility that the intermediate influences CRP and cancer through additional cell types or tissues not included in this analysis. Third, we cannot rule out the possibility that candidate causal variants at multi-trait colocalised loci influenced CRP and cancer through biological mechanisms independent to the molecular intermediate identified (e.g. through co-regulation of a neighbouring gene in a tissue that was not evaluated)^87^. Fourth, we did not examine anatomical subsite or histological subtype-stratified cancer outcomes. Finally, our analyses were restricted to participants of European ancestry and therefore have unclear generalisability to other non-European populations.

Our findings motivate several future lines of research. First, follow-up of molecular intermediates identified in our work by exploring the contribution of rare variants in genes encoding these intermediates on cancer risk could provide orthogonal evidence into their effects. Second, our findings implicating targets of approved medications in cancer risk warrant further investigation using electronic health record data to validate their repurposing potential and preclinical models to clarify potential mechanisms of effect. In addition, extending the approach performed here to other ancestries can enable exploration of whether our findings are transportable to non-European populations. By anchoring non-causal biomarker-disease associations to their upstream genetic and molecular determinants, our framework enables the discovery of causal mechanisms contributing to disease onset and the prioritisation of novel therapeutic targets for disease prevention which we have illustrated through application to CRP and cancer. We envision that this approach could have application for uncovering potential phenotypic confounders underlying epidemiological relationships beyond CRP and cancer.

## Methods

### Observational analysis of pre-diagnostic CRP concentrations and cancer risk in the UK Biobank

The UK Biobank is a prospective cohort study of 502,507 participants from England, Scotland, and Wales who were recruited between 2006 and 2010^88^. All participants completed a series of physical, sociodemographic, and medical assessments and provided blood, urine, and saliva samples at baseline. All participants provided written informed consent prior to data collection.

In our analyses we excluded participants with a cancer diagnosis prior to baseline assessment (N=46,486), those with a diagnosis of a cancer type not included in the 11 cancer outcomes (N= 41,416), and those with missing data on CRP or covariates (N=15,211-46,623 depending on the cancer endpoint and model), leaving an analytic cohort of 414,034 participants.

Serum CRP was measured at baseline by immunoturbidimetric high-sensitivity analysis on a Beckman Coulter AU5800 and natural log-transformed prior to analysis. We selected 11 cancer outcomes that have previously been linked to CRP in observational studies where large-scale genetic association data on these outcomes were also available. Cancer outcomes were defined using ICD-10 codes and incident cases were derived from national cancer registries by taking the first diagnosed cancer per participant. Person-years were calculated from the UK Biobank baseline assessment date to the date of first registration of cancer diagnosis, death, lost to follow-up, or the end of follow-up, whichever came first. The end of follow-up date was assigned based on the cancer registry censoring date indicated by UK Biobank, which was 31 December 2020 for England, 31 December 2016 for Wales, and 30 November 2021 for Scotland; for each participant, the country was inferred from the UK Biobank assessment centre attended at baseline assessment.

We employed Cox proportional hazards models to estimate hazard ratios (HRs) and 95% confidence intervals (CIs) for the association between baseline circulating CRP concentrations and cancer risk. We considered both a “minimally adjusted” model with sex, age at recruitment, and ethnicity as covariates and a “fully adjusted” model that additionally included socioeconomic factors (education, Townsend deprivation index), anthropometric traits (body mass index, BMI; height), lifestyle factors (smoking status, alcohol use, physical activity), and family history of cancer. In female-specific cancers (i.e. breast, endometrial, ovarian), these models were further adjusted for menopausal status, oral contraceptive use, and hormone replacement therapy use. To evaluate if findings were influenced by reverse causation, lag analyses were performed by excluding participants within the first 3 years of follow-up. Further information on cancer case definition and covariate classification is presented in the **Supplementary Materials**.

### Evaluating causal relationships between plasma CRP and cancer risk using bidirectional MR

Summary genetic association data on plasma CRP were obtained from a GWAS performed in 575,531 participants of European ancestry in the Cohorts for Heart and Aging Research in Genomic Epidemiology (CHARGE) Consortium^89^. Summary genetic association data were also obtained from GWAS of 10 cancer outcomes (breast, colorectal, endometrial, esophageal, head and neck, lung, non-Hodgkin lymphoma, pancreatic, prostate, renal) in 431,213 cases and up to 2,207,505 controls of European ancestry^90-99^. A summary of case and control numbers across each cancer outcome is presented in **Table S1**. We also obtained summary genetic association data on a lifetime smoking index (defined using information on smoking duration, heaviness, and cessation) from 462,690 individuals in the UK Biobank to perform multivariable MR analysis of the association of genetic liability to CRP on lung cancer risk adjusted for this index^100^. All studies contributing data to these analyses had the relevant institutional review board approval from each country, in accordance with the Declaration of Helsinki, and all participants provided informed consent.

Mendelian randomization assumes that a genetic instrument i) is associated with the exposure (“relevance”), ii) has no common causes with an outcome (“exchangeability”), and iii) does not have a direct effect on an outcome (“exclusion restriction”)^7^. We tested the “relevance” assumption by calculating F-statistics for genetic instruments^101^. The restriction of participants across GWAS to individuals of primarily European ancestry and the adjustment for principal components of genetic ancestry within studies should minimise violations of the “exchangeability” assumption through confounding by population stratification. In addition, this should ensure that participants in these studies are representative of the same underlying population. We evaluated the robustness of findings to violations of the “exclusion restriction” assumption through use of various pleiotropy-robust models (described below).

To evaluate the effect of circulating CRP on cancer risk (forward MR), we constructed a genetic instrument for circulating CRP levels from genome-wide significant (*P*<5x10^-8^) and independent (LD r^2^<0.001) SNPs located in or within 100 kb from the *CRP* gene using a reference panel of 10,000 randomly selected and unrelated UK Biobank participants of white British ancestry. To evaluate the effect of genetic liability to cancer on circulating CRP levels (reverse MR), we constructed genetic instruments for each cancer outcome by identifying SNPs associated with susceptibility to that cancer (*P*<5x10^-8^, LD r^2^<0.001). Characteristics of genetic variants used as instruments to proxy circulating CRP and liability to site-specific cancers are presented in **ST20**.

In both forward (CRP → cancer) and reverse (cancer → CRP) MR analyses, we used inverse-variance weighted random effects models to estimate causal effects. In reverse MR analyses, we also employed four “pleiotropy-robust” models to evaluate whether associations were influenced by horizontal pleiotropy: MR-Egger regression, weighted median estimation, CAUSE, and MR-CUE^102-105^. Two of these methods can produce biased causal estimates in the presence of correlated pleiotropy (i.e. MR-Egger, weighted median) whereas CAUSE and MR-CUE account for this form of pleiotropy. In CAUSE, we compared if a “causal” model (permitting both a causal effect and horizontal pleiotropy) fit our data better than a “sharing” model (permitting horizontal pleiotropy only) using the expected log pointwise posterior density (ELPD). We also performed multivariable Mendelian randomization to adjust for potential sources of correlated pleiotropy using MRBEE (MR using bias-corrected estimating equation) which is more robust to weak instruments^106^.

MR analyses were performed using the TwoSampleMR R package using R version 4.6.1 and were not pre-registered.

### Cross-trait pleiotropy analysis applied to CRP and cancer

To identify pleiotropic loci that influence CRP and cancer we used pleiotropic analysis under a composite null hypothesis (PLACO) in pairwise analyses^44^. In brief, PLACO tests the composite null hypothesis that a variant is associated with none or only 1 of 2 traits. The test statistic is generated as the product of the Z-statistic of the association of each variant with each trait (*Z*^2^), which is assumed to follow a mixture distribution. We removed variants with a minor allele frequency (MAF) < 0.01 prior to performing PLACO. We performed decorrelation of Z-score matrices to account for modest sample overlap across select studies. For all variants with strong evidence of having pleiotropic effects on CRP and cancer (*P*_PLACO_<5×10^-8^), we then identified distinct pleiotropic loci by clumping variants using LD thresholds set at r^2^=0.6 (to determine the coordinates of each locus) and r^2^=0.1 (to define independent signals) using the 1000 genomes phase 3 reference panel into a single genetic locus using FUMA (SNP2GENE function, v1.3.6a).^107^ All SNPs in LD with each other at 0.01≥r^2^<0.6 were assigned to the same LD block. LD blocks were merged into one locus if there were SNPs from different LD blocks closer than 250 kb.

### CRP and cancer colocalisation analysis

Colocalisation was performed to provide evidence of shared causal variants at pleiotropic loci associated with CRP and cancer that were identified in PLACO^108^. We used the coloc package to generate posterior probabilities (PPs) that associations between CRP and cancer represent each of the following configurations: (1) neither CRP nor cancer has a genetic association in the region (H_0_), (2) only CRP has a genetic association in the region (H_1_), (3) only cancer has a genetic association in the region (H_2_), (4) both CRP and cancer have a genetic association in the region but have different causal variants (H_3_), and (5) both CRP and cancer have a shared causal variant in the region (H_4_). Colocalisation was performed by generating ±100 kb windows around the lead SNP in each pleiotropic locus. We only proceeded with colocalisation analyses where >150 SNPs were present in a genomic window. We employed default priors for p1 (ie, prior probability that a variant is associated with CRP, 1x10^-4^), p2 (ie, prior probability that a variant is associated with cancer, 1x10^-4^), and p12 (ie, prior probability that a variant is associated with both traits, 1x10^-5^). We used a posterior probability of colocalisation (PPH_4_)>50% to indicate support for colocalisation of CRP and cancer associations at each locus, as this represents the posterior probability with the majority support across all configurations tested. For all pleiotropic loci with evidence of colocalisation, we identified the variant with the highest posterior probability of being the causal variant.

### Mapping of colocalised pleiotropic loci to genes and identification of novel susceptibility loci

To identify candidate effector genes at colocalised pleiotropic loci, we then mapped the putative causal variant at each locus to the nearest protein-coding gene using Ensembl Biomart Build 37. We queried all genes where the start and end coordinates are ±500 kb from the lead SNP. The gene with the smallest interval between each SNP and the gene’s interval is reported as the nearest protein-coding gene. For all candidate effector genes mapped to colocalised pleiotropic loci, we explored if loci in or in proximity to these genes have previously been mapped to site-specific cancer using LDlinkR. We defined loci as novel cancer susceptibility loci if there was no SNP within ± 500 kb or LD *r*^2^>0.10 from the lead SNP in the locus previously reported to be associated with the relevant cancer outcome at *P*<5x10^-8^ in the GWAS Catalog (as of 3 July 2026).

### Identifying molecular phenotypes that mediate effects at colocalised loci

At each pleiotropic locus with evidence of colocalisation, we then investigated molecular phenotypes that mediate these effects by integrating molecular QTL data into HyprColoc multi-trait colocalisation. Specifically, we used genetic data on plasma protein levels from previous analyses of the UK Biobank (N_proteins_=2,923, N_participants_=54,219) and deCODE (N_proteins_=4,907, N_participants_=35,559) studies, and whole blood gene expression from the eQTLGen consortium (N=31,684)^109-111^. In analyses examining immune cell single-cell gene expression, we also obtained data from peripheral blood mononuclear cells (PBMCs) on 14 immune cells (plasma cells, immature and naive B cells, memory B cells, naive and central memory T [CD4_NC_], effector memory and central memory T [CD4_ET_], SOX4-expressing T [CD4_SOX4_] cells, CD8_NC_, CD8_ET_, CD8_SOX4_ cells, natural killer [NK] and NK-recruiting cells, classical [Mono_C_], non-classical [Mono_NC_] monocytes, dendritic cells [DCs]) in the OneK1K study (N=982) and CD4^+^ memory and naïve T cells that were either in a resting state (i.e. unstimulated) or stimulated at one of three time points (16h, 40h, 5 days) in Soskic *et al* (N=119)^14,112,113^. All studies were performed in individuals of European ancestry. Further information on the molecular traits included from each study is presented in the **Supplementary Materials**. We restricted analyses to pleiotropic loci where the lead SNP was within 100 kb from a *cis*-acting SNP that influenced protein or gene expression of a neighbouring gene (*P*<5x10^-6^). HyprColoc was then performed on a genomic window ±100 kb around the lead SNP at each pleiotropic locus using default variant-specific prior configuration; priors 1 and 2 were set at 1x10^−4^ and 0.02, respectively; and regional and alignment thresholds of 0.5 were used. We used a PPH_4_>50% to indicate support for colocalisation of CRP, cancer, and putative molecular intermediates at each locus. We declared a molecular phenotype with multi-trait colocalisation evidence at a particular locus as a candidate molecular intermediate at that locus where the most likely causal variant was not in LD (r^2^<0.05, 1000 Genomes Phase 3 CEU panel) with a candidate causal variant underlying a multi-trait colocalised signal for the same cancer type and a different molecular phenotype.

### Evaluation of pleiotropy through causal co-regulation of neighbouring genes

Given evidence of widespread co-regulation of eQTL with neighbouring genes, we further evaluated if findings at multi-trait colocalised loci were potentially biased by pleiotropy via other molecular phenotypes by examining if the putative causal variant at each locus, or a SNP in LD with this variant (LD r^2^>0.10 using the 1000 Genomes Phase 3 CEU reference panel), causally influenced two or more candidate molecular phenotypes (i.e. PPH_4_ > 50%)^87^. Where two or more molecular phenotypes with multi-trait colocalisation evidence were identified at the same locus, we de-prioritised further characterisation of these loci as we were unable to confidently assign a single molecular trait as a likely mediator of the shared genetic signal identified.

### Examining drug repurposing and druggability of candidate effector genes

We searched ChEMBL and Therapeutic Target Database to identify approved or investigational medications targeting the products of candidate effector genes and molecular intermediates to inform on their potential repurposing potential^114,115^. We also investigated the druggability of candidate effector genes mapped to colocalised pleiotropic loci using data from Finan *et al*^116^. This analysis combined data on protein targets of known and investigational medications, proteins with sequence similarities to these targets, and secreted or extracellular proteins belonging to druggable families, to identify 4,479 genes that are drugged or considered druggable.

## Data availability

This research has been conducted using the UK Biobank Resource under Application Number 22102. Applications to access the data from bone fide researchers can be made at https://www.ukbiobank.ac.uk/enable-your-research/apply-for-access. Summary genetic association data on circulating CRP and select site-specific cancers can be obtained from the GWAS catalog using the following study accession IDs: CRP (GCST90029070), colorectal cancer (GCST90255675), esophageal cancer (GCST003739), lung cancer (GCST004748), endometrial cancer (GCST006464), renal cancer (GCST90320057), non-Hodgkin lymphoma (GCST90011819), prostate cancer (GCST90274714), and breast cancer (GCST010098). Head and neck cancer data can be obtained from the MRC IEU OpenGWAS database under accession number ieu-b-5129 (https://opengwas.io/datasets/ieu-b-5129). Pancreatic cancer data (phs000206.v7.p3, phs000648.v2.p1) were obtained from dbGap applications 148652-1 and 148653-1. Lifetime smoking GWAS data were downloaded from https://doi.org/10.5523/bris.10i96zb8gm0j81yz0q6ztei23d. Plasma proteomics data from Sun et al. (Nature, 2023) were downloaded from https://metabolomics.helmholtz-munich.de/ukbbpgwas/. Plasma proteomics data from Ferkingstad et al. (Nature Genetics, 2021) were downloaded from https://www.decode.com/summarydata/. Bulk blood gene expression data were downloaded from https://www.eqtlgen.org/. Single-cell immune cell gene expression data from Yazar et al. (Science, 2022) were downloaded from https://onek1k.org/. Single-cell immune cell gene expression data from Soskic et al. (Nature Genetics, 2022) were downloaded from https://trynkalab.sanger.ac.uk.

## IARC Disclaimer

Where authors are identified as personnel of the International Agency for Research on Cancer/World Health Organization, the authors alone are responsible for the views expressed in this article and they do not necessarily represent the decisions, policy, or views of the International Agency for Research on Cancer/World Health Organization.

## References

1 Rothman, K. J., Greenland, S. & Lash, T. L. Modern epidemiology. (Philadelphia : Wolters Kluwer Health/Lippincott Williams & Wilkins, 2008).

2 Tennant, P. W. G. et al. Use of directed acyclic graphs (DAGs) to identify confounders in applied health research: review and recommendations. Int J Epidemiol 50, 620–632 (2021). 10.1093/ije/dyaa213

3 Hammerton, G. & Munafò, M. R. Causal inference with observational data: the need for triangulation of evidence. Psychol Med 51, 563–578 (2021). 10.1017/s0033291720005127

4 Fewell, Z., Davey Smith, G. & Sterne, J. A. The impact of residual and unmeasured confounding in epidemiologic studies: a simulation study. Am J Epidemiol 166, 646–655 (2007). 10.1093/aje/kwm165

5 Phillips, A. N. & Smith, G. D. How independent are "independent" effects? Relative risk estimation when correlated exposures are measured imprecisely. J Clin Epidemiol 44, 1223–1231 (1991). 10.1016/0895-4356(91)90155-3

6 Pingault, J. B., Richmond, R. & Davey Smith, G. Causal Inference with Genetic Data: Past, Present, and Future. Cold Spring Harb Perspect Med 12 (2022). 10.1101/cshperspect.a041271

7 Davey Smith, G. & Hemani, G. Mendelian randomization: genetic anchors for causal inference in epidemiological studies. Hum Mol Genet 23, R89–98 (2014). 10.1093/hmg/ddu328

8 Trajanoska, K. et al. From target discovery to clinical drug development with human genetics. Nature 620, 737–745 (2023). 10.1038/s41586-023-06388-8

9 Smith, G. D. & Ebrahim, S. ’Mendelian randomization’: can genetic epidemiology contribute to understanding environmental determinants of disease? Int J Epidemiol 32, 1–22 (2003). 10.1093/ije/dyg070

10 Smith, G. D. et al. Clustered environments and randomized genes: a fundamental distinction between conventional and genetic epidemiology. PLoS Med 4, e352 (2007). 10.1371/journal.pmed.0040352

11 Davies, N. M., Holmes, M. V. & Davey Smith, G. Reading Mendelian randomisation studies: a guide, glossary, and checklist for clinicians. Bmj 362, k601 (2018). 10.1136/bmj.k601

12 Minikel, E. V., Painter, J. L., Dong, C. C. & Nelson, M. R. Refining the impact of genetic evidence on clinical success. Nature 629, 624–629 (2024). 10.1038/s41586-024-07316-0

13 Burgess, S. et al. Using genetic association data to guide drug discovery and development: Review of methods and applications. Am J Hum Genet 110, 195–214 (2023). 10.1016/j.ajhg.2022.12.017

14 The blood proteome of imminent lung cancer diagnosis. Nat Commun 14, 3042 (2023). 10.1038/s41467-023-37979-8

15 Pepys, M. B. & Hirschfield, G. M. C-reactive protein: a critical update. J Clin Invest 111, 1805–1812 (2003). 10.1172/jci18921

16 Sproston, N. R. & Ashworth, J. J. Role of C-Reactive Protein at Sites of Inflammation and Infection. Front Immunol 9, 754 (2018). 10.3389/fimmu.2018.00754

17 Allin, K. H., Bojesen, S. E. & Nordestgaard, B. G. Baseline C-reactive protein is associated with incident cancer and survival in patients with cancer. J Clin Oncol 27, 2217–2224 (2009). 10.1200/jco.2008.19.8440

18 Erlinger, T. P., Platz, E. A., Rifai, N. & Helzlsouer, K. J. C-reactive protein and the risk of incident colorectal cancer. Jama 291, 585–590 (2004). 10.1001/jama.291.5.585

19 Gunter, M. J. et al. A prospective study of serum C-reactive protein and colorectal cancer risk in men. Cancer Res 66, 2483–2487 (2006). 10.1158/0008-5472.Can-05-3631

20 Gunter, M. J. et al. Circulating Adipokines and Inflammatory Markers and Postmenopausal Breast Cancer Risk. J Natl Cancer Inst 107 (2015). 10.1093/jnci/djv169

21 Il’yasova, D. et al. Circulating levels of inflammatory markers and cancer risk in the health aging and body composition cohort. Cancer Epidemiol Biomarkers Prev 14, 2413–2418 (2005). 10.1158/1055-9965.Epi-05-0316

22 Otani, T., Iwasaki, M., Sasazuki, S., Inoue, M. & Tsugane, S. Plasma C-reactive protein and risk of colorectal cancer in a nested case-control study: Japan Public Health Center-based prospective study. Cancer Epidemiol Biomarkers Prev 15, 690–695 (2006). 10.1158/1055-9965.Epi-05-0708

23 Peres, L. C. et al. High Levels of C-Reactive Protein Are Associated with an Increased Risk of Ovarian Cancer: Results from the Ovarian Cancer Cohort Consortium. Cancer Res 79, 5442–5451 (2019). 10.1158/0008-5472.Can-19-1554

24 Pletnikoff, P. P. et al. Cardiorespiratory fitness, C-reactive protein and lung cancer risk: A prospective population-based cohort study. Eur J Cancer 51, 1365–1370 (2015). 10.1016/j.ejca.2015.04.020

25 Trichopoulos, D., Psaltopoulou, T., Orfanos, P., Trichopoulou, A. & Boffetta, P. Plasma C-reactive protein and risk of cancer: a prospective study from Greece. Cancer Epidemiol Biomarkers Prev 15, 381–384 (2006). 10.1158/1055-9965.Epi-05-0626

26 Tsilidis, K. K. et al. C-reactive protein and colorectal cancer risk: a systematic review of prospective studies. Int J Cancer 123, 1133–1140 (2008). 10.1002/ijc.23606

27 Allin, K. H., Nordestgaard, B. G., Zacho, J., Tybjaerg-Hansen, A. & Bojesen, S. E. C-reactive protein and the risk of cancer: a mendelian randomization study. J Natl Cancer Inst 102, 202–206 (2010). 10.1093/jnci/djp459

28 He, C. et al. Genetically Predicted Circulating Level of C-Reactive Protein Is Not Associated With Prostate Cancer Risk. Front Oncol 10, 545603 (2020). 10.3389/fonc.2020.545603

29 Heikkilä, K. et al. C-reactive protein-associated genetic variants and cancer risk: findings from FINRISK 1992, FINRISK 1997 and Health 2000 studies. Eur J Cancer 47, 404-412 (2011). 10.1016/j.ejca.2010.07.032

30 Ji, M. et al. Circulating C-reactive protein increases lung cancer risk: Results from a prospective cohort of UK Biobank. Int J Cancer 150, 47–55 (2022). 10.1002/ijc.33780

31 Robinson, T., Martin, R. M. & Yarmolinsky, J. Mendelian randomisation analysis of circulating adipokines and C-reactive protein on breast cancer risk. Int J Cancer 147, 1597–1603 (2020). 10.1002/ijc.32947

32 Wang, X. et al. Mendelian randomization analysis of C-reactive protein on colorectal cancer risk. Int J Epidemiol 48, 767–780 (2019). 10.1093/ije/dyy244

33 Davey Smith, G., Hemani, G. & Ebrahim, S. Gene-environment equivalence: The fundamental principle of Mendelian randomization. PLoS Med 23, e1005013 (2026). 10.1371/journal.pmed.1005013

34 Ding, M., Bhupathiraju, S. N., Satija, A., van Dam, R. M. & Hu, F. B. Long-term coffee consumption and risk of cardiovascular disease: a systematic review and a dose-response meta-analysis of prospective cohort studies. Circulation 129, 643–659 (2014). 10.1161/circulationaha.113.005925

35 Klatsky, A. L., Koplik, S., Kipp, H. & Friedman, G. D. The confounded relation of coffee drinking to coronary artery disease. Am J Cardiol 101, 825–827 (2008). 10.1016/j.amjcard.2007.11.022

36 Kleemola, P., Jousilahti, P., Pietinen, P., Vartiainen, E. & Tuomilehto, J. Coffee consumption and the risk of coronary heart disease and death. Arch Intern Med 160, 3393–3400 (2000). 10.1001/archinte.160.22.3393

37 Lopez-Garcia, E. et al. Coffee consumption and coronary heart disease in men and women: a prospective cohort study. Circulation 113, 2045–2053 (2006). 10.1161/circulationaha.105.598664

38 Liu, M. et al. Association studies of up to 1.2 million individuals yield new insights into the genetic etiology of tobacco and alcohol use. Nat Genet 51, 237–244 (2019). 10.1038/s41588-018-0307-5

39 Lima de Castro, F. B. A. et al. Acute Effects of Coffee Consumption on Blood Pressure and Endothelial Function in Individuals with Hypertension on Antihypertensive Drug Treatment: A Randomized Crossover Trial. High Blood Press Cardiovasc Prev 31, 65-76 (2024). 10.1007/s40292-024-00622-8

40 Marcus, G. M. et al. Acute Effects of Coffee Consumption on Health among Ambulatory Adults. N Engl J Med 388, 1092–1100 (2023). 10.1056/NEJMoa2204737

41 Nijssen, K. M. R., Mensink, R. P. & Joris, P. J. Effects of Coffee-Related Bioactive Components on Flow-Mediated Vasodilation: A Meta-Analysis of Randomized, Controlled Intervention Studies in Adults. Nutr Rev (2025). 10.1093/nutrit/nuaf211

42 Nordestgaard, A. T. & Nordestgaard, B. G. Coffee intake, cardiovascular disease and all-cause mortality: observational and Mendelian randomization analyses in 95 000-223 000 individuals. Int J Epidemiol 45, 1938–1952 (2016). 10.1093/ije/dyw325

43 Zhu, M. et al. C-reactive protein and cancer risk: a pan-cancer study of prospective cohort and Mendelian randomization analysis. BMC Med 20, 301 (2022). 10.1186/s12916-022-02506-x

44 Ray, D. & Chatterjee, N. A powerful method for pleiotropic analysis under composite null hypothesis identifies novel shared loci between Type 2 Diabetes and Prostate Cancer. PLoS Genet 16, e1009218 (2020). 10.1371/journal.pgen.1009218

45 Doll, R. & Hill, A. B. Smoking and carcinoma of the lung; preliminary report. Br Med J 2, 739–748 (1950). 10.1136/bmj.2.4682.739

46 Lassi, G. et al. The CHRNA5-A3-B4 Gene Cluster and Smoking: From Discovery to Therapeutics. Trends Neurosci 39, 851–861 (2016). 10.1016/j.tins.2016.10.005

47 Galan, D. et al. Applying Mendelian randomization to appraise causality in relationships between smoking, depression and inflammation. Sci Rep 12, 15041 (2022). 10.1038/s41598-022-19214-4

48 Buchwald, J. et al. Genome-wide association meta-analysis of nicotine metabolism and cigarette consumption measures in smokers of European descent. Mol Psychiatry 26, 2212–2223 (2021). 10.1038/s41380-020-0702-z

49 Di Credico, G. et al. Alcohol drinking and head and neck cancer risk: the joint effect of intensity and duration. Br J Cancer 123, 1456–1463 (2020). 10.1038/s41416-020-01031-z

50 Holmes, M. V. et al. Association between alcohol and cardiovascular disease: Mendelian randomisation analysis based on individual participant data. Bmj 349, g4164 (2014). 10.1136/bmj.g4164

51 Kranzler, H. R. et al. Genome-wide association study of alcohol consumption and use disorder in 274,424 individuals from multiple populations. Nat Commun 10, 1499 (2019). 10.1038/s41467-019-09480-8

52 Ter Steege, E. J., et al. R-spondin-3 is an oncogenic driver of poorly differentiated invasive breast cancer. J Pathol 258, 289–299 (2022). 10.1002/path.5999

53 Ter Steege, E. J., et al. R-spondin-3 promotes proliferation and invasion of breast cancer cells independently of Wnt signaling. Cancer Lett 568, 216301 (2023). 10.1016/j.canlet.2023.216301

54 Feng, Y. et al. LRRC25 Functions as an Inhibitor of NF-κB Signaling Pathway by Promoting p65/RelA for Autophagic Degradation. Sci Rep 7, 13448 (2017). 10.1038/s41598-017-12573-3

55 Zhang, G., Yu, H., Liu, J., Dong, G. & Cai, Z. Myeloid-lineage-specific membrane protein LRRC25 suppresses immunity in solid tumor and is a potential cancer immunotherapy checkpoint target. Cell Rep 44, 115631 (2025). 10.1016/j.celrep.2025.115631

56 Abel, E. V. et al. HNF1A is a novel oncogene that regulates human pancreatic cancer stem cell properties. Elife 7 (2018). 10.7554/eLife.33947

57 Cheng, K. et al. Specific activation of the TLR1-TLR2 heterodimer by small-molecule agonists. Sci Adv 1 (2015). 10.1126/sciadv.1400139

58 Kishore, M. et al. Regulatory T Cell Migration Is Dependent on Glucokinase-Mediated Glycolysis. Immunity 47, 875–889.e810 (2017). 10.1016/j.immuni.2017.10.017

59 Xie, Z., Xie, T., Liu, J., Zhang, Q. & Xiao, X. Glucokinase Inactivation Ameliorates Lipid Accumulation and Exerts Favorable Effects on Lipid Metabolism in Hepatocytes. Int J Mol Sci 24 (2023). 10.3390/ijms24054315

60 Zhu, D. et al. Dorzagliatin in drug-naïve patients with type 2 diabetes: a randomized, double-blind, placebo-controlled phase 3 trial. Nat Med 28, 965–973 (2022). 10.1038/s41591-022-01802-6

61 Yang, H. & Geiger, M. Cell penetrating SERPINA5 (ProteinC inhibitor, PCI): More questions than answers. Semin Cell Dev Biol 62, 187–193 (2017). 10.1016/j.semcdb.2016.10.007

62 Fan, M. et al. SERPINA5 promotes tumour cell proliferation by modulating the PI3K/AKT/mTOR signalling pathway in gastric cancer. J Cell Mol Med 26, 4837–4846 (2022). 10.1111/jcmm.17514

63 MacLeod, M. K., Clambey, E. T., Kappler, J. W. & Marrack, P. CD4 memory T cells: what are they and what can they do? Semin Immunol 21, 53–61 (2009). 10.1016/j.smim.2009.02.006

64 Couzinet, A. et al. A cell-type-specific requirement for IFN regulatory factor 5 (IRF5) in Fas-induced apoptosis. Proc Natl Acad Sci U S A 105, 2556–2561 (2008). 10.1073/pnas.0712295105

65 Yan, J., Pandey, S. P., Barnes, B. J., Turner, J. R. & Abraham, C. T Cell-Intrinsic IRF5 Regulates T Cell Signaling, Migration, and Differentiation and Promotes Intestinal Inflammation. Cell Rep 31, 107820 (2020). 10.1016/j.celrep.2020.107820

66 Xiao, J. et al. L3MBTL3 and STAT3 collaboratively upregulate SNAIL expression to promote metastasis in female breast cancer. Nat Commun 16, 231 (2025). 10.1038/s41467-024-55617-9

67 Papp, K. et al. Apremilast, an oral phosphodiesterase 4 (PDE4) inhibitor, in patients with moderate to severe plaque psoriasis: Results of a phase III, randomized, controlled trial (Efficacy and Safety Trial Evaluating the Effects of Apremilast in Psoriasis [ESTEEM] 1). J Am Acad Dermatol 73, 37–49 (2015). 10.1016/j.jaad.2015.03.049

68 Genovese, M. C. et al. Apremilast in Patients With Active Rheumatoid Arthritis: A Phase II, Multicenter, Randomized, Double-Blind, Placebo-Controlled, Parallel-Group Study. Arthritis Rheumatol 67, 1703–1710 (2015). 10.1002/art.39120

69 Ram, T. et al. MEK inhibitors in cancer treatment: structural insights, regulation, recent advances and future perspectives. RSC Med Chem 14, 1837–1857 (2023). 10.1039/d3md00145h

70 Zhang, J. & Chen, Z. Integration of TWAS with single-cell and spatial transcriptomics identifies TLR1 as a susceptibility gene and therapeutic target in the breast cancer tumor microenvironment. Int J Biol Macromol 353, 150951 (2026). 10.1016/j.ijbiomac.2026.150951

71 Mukherjee, S. et al. Toll-like receptor-guided therapeutic intervention of human cancers: molecular and immunological perspectives. Front Immunol 14, 1244345 (2023). 10.3389/fimmu.2023.1244345

72 Liu, W. et al. LRRC25 plays a key role in all-trans retinoic acid-induced granulocytic differentiation as a novel potential leukocyte differentiation antigen. Protein Cell 9, 785–798 (2018). 10.1007/s13238-017-0421-7

73 Weiss, M., Blazek, K., Byrne, A. J., Perocheau, D. P. & Udalova, I. A. IRF5 is a specific marker of inflammatory macrophages in vivo. Mediators Inflamm 2013, 245804 (2013). 10.1155/2013/245804

74 Roberts, B. K., Collado, G. & Barnes, B. J. Role of interferon regulatory factor 5 (IRF5) in tumor progression: Prognostic and therapeutic potential. Biochim Biophys Acta Rev Cancer 1879, 189061 (2024). 10.1016/j.bbcan.2023.189061

75 Harrison, S. A. et al. A randomized, placebo-controlled trial of emricasan in patients with NASH and F1-F3 fibrosis. J Hepatol 72, 816–827 (2020). 10.1016/j.jhep.2019.11.024

76 Iqbal, L., Zameer, U. & Iqbal Malick, M. Exploring Talarozole as a Novel Therapeutic Approach for Osteoarthritis: Insights From Experimental Studies. Clin Med Insights Arthritis Musculoskelet Disord 17, 11795441231222494 (2024). 10.1177/11795441231222494

77 Ridker, P. M. et al. IL-6 inhibition with ziltivekimab in patients at high atherosclerotic risk (RESCUE): a double-blind, randomised, placebo-controlled, phase 2 trial. Lancet 397, 2060–2069 (2021). 10.1016/s0140-6736(21)00520-1

78 Beck, T. C. et al. Cellular and Molecular Mechanisms of MEK1 Inhibitor-Induced Cardiotoxicity. JACC CardioOncol 4, 535–548 (2022). 10.1016/j.jaccao.2022.07.009

79 Schett, G., Sloan, V. S., Stevens, R. M. & Schafer, P. Apremilast: a novel PDE4 inhibitor in the treatment of autoimmune and inflammatory diseases. Ther Adv Musculoskelet Dis 2, 271–278 (2010). 10.1177/1759720x10381432

80 Ridker, P. M. et al. Effect of interleukin-1β inhibition with canakinumab on incident lung cancer in patients with atherosclerosis: exploratory results from a randomised, double-blind, placebo-controlled trial. Lancet 390, 1833–1842 (2017). 10.1016/s0140-6736(17)32247-x

81 Novo Nordisk provides update on the ZEUS phase 3 trial in people with ASCVD, CKD and inflammation, <https://www.novonordisk.com/content/nncorp/global/en/news-and-media/news-and-ir-materials/news-details.html?id=916587&_sp=1792bd3a-9dd2-4522-902d-a55b275b7bfe.1785767208184> (2026).

82 Chertow, G. M. et al. IL-6 inhibition with clazakizumab in patients receiving maintenance dialysis: a randomized phase 2b trial. Nat Med 30, 2328–2336 (2024). 10.1038/s41591-024-03043-1

83 Sanderson, E. et al. Heritable confounding in Mendelian randomization studies: structure, consequences and relevance for gene-environment equivalence. medRxiv, 2024.2009.2005.24312293 (2025). 10.1101/2024.09.05.24312293

84 Warwick, A. N. et al. Harnessing confounding and genetic pleiotropy to identify causes of disease through proteomics and Mendelian randomisation – ‘MR Fish’. medRxiv, 2024.2007.2011.24310200 (2024). 10.1101/2024.07.11.24310200

85 Hamilton, F., Pedersen, K. M., Ghazal, P., Nordestgaard, B. G. & Smith, G. D. Low levels of small HDL particles predict but do not influence risk of sepsis. Crit Care 27, 389 (2023). 10.1186/s13054-023-04589-1

86 Visscher, P. M., Yengo, L., Cox, N. J. & Wray, N. R. Discovery and implications of polygenicity of common diseases. Science 373, 1468–1473 (2021). 10.1126/science.abi8206

87 Tambets, R., Kolde, A., Kolberg, P., Love, M. I. & Alasoo, K. Extensive co-regulation of neighboring genes complicates the use of eQTLs in target gene prioritization. HGG Adv 5, 100348 (2024). 10.1016/j.xhgg.2024.100348

88 Sudlow, C. et al. UK biobank: an open access resource for identifying the causes of a wide range of complex diseases of middle and old age. PLoS Med 12, e1001779 (2015). 10.1371/journal.pmed.1001779

89 Said, S. et al. Genetic analysis of over half a million people characterises C-reactive protein loci. Nat Commun 13, 2198 (2022). 10.1038/s41467-022-29650-5

90 Ebrahimi, E. et al. Cross-ancestral GWAS identifies 29 variants across head and neck cancer subsites. Nat Commun 16, 8787 (2025). 10.1038/s41467-025-63842-z

91 Fernandez-Rozadilla, C. et al. Deciphering colorectal cancer genetics through multi-omic analysis of 100,204 cases and 154,587 controls of European and east Asian ancestries. Nat Genet 55, 89–99 (2023). 10.1038/s41588-022-01222-9

92 Gharahkhani, P. et al. Genome-wide association studies in oesophageal adenocarcinoma and Barrett’s oesophagus: a large-scale meta-analysis. Lancet Oncol 17, 1363–1373 (2016). 10.1016/s1470-2045(16)30240-6

93 Klein, A. P. et al. Genome-wide meta-analysis identifies five new susceptibility loci for pancreatic cancer. Nat Commun 9, 556 (2018). 10.1038/s41467-018-02942-5

94 McKay, J. D. et al. Large-scale association analysis identifies new lung cancer susceptibility loci and heterogeneity in genetic susceptibility across histological subtypes. Nat Genet 49, 1126–1132 (2017). 10.1038/ng.3892

95 O’Mara, T. A. et al. Identification of nine new susceptibility loci for endometrial cancer. Nat Commun 9, 3166 (2018). 10.1038/s41467-018-05427-7

96 Purdue, M. P. et al. Multi-ancestry genome-wide association study of kidney cancer identifies 63 susceptibility regions. Nat Genet 56, 809–818 (2024). 10.1038/s41588-024-01725-7

97 Rashkin, S. R. et al. Pan-cancer study detects genetic risk variants and shared genetic basis in two large cohorts. Nat Commun 11, 4423 (2020). 10.1038/s41467-020-18246-6

98 Wang, A. et al. Characterizing prostate cancer risk through multi-ancestry genome-wide discovery of 187 novel risk variants. Nat Genet 55, 2065–2074 (2023). 10.1038/s41588-023-01534-4

99 Zhang, H. et al. Genome-wide association study identifies 32 novel breast cancer susceptibility loci from overall and subtype-specific analyses. Nat Genet 52, 572–581 (2020). 10.1038/s41588-020-0609-2

100 Wootton, R. E. et al. Evidence for causal effects of lifetime smoking on risk for depression and schizophrenia: a Mendelian randomisation study. Psychol Med 50, 2435–2443 (2020). 10.1017/s0033291719002678

101 Burgess, S. & Thompson, S. G. Avoiding bias from weak instruments in Mendelian randomization studies. Int J Epidemiol 40, 755–764 (2011). 10.1093/ije/dyr036

102 Bowden, J., Davey Smith, G. & Burgess, S. Mendelian randomization with invalid instruments: effect estimation and bias detection through Egger regression. Int J Epidemiol 44, 512–525 (2015). 10.1093/ije/dyv080

103 Bowden, J., Davey Smith, G., Haycock, P. C. & Burgess, S. Consistent Estimation in Mendelian Randomization with Some Invalid Instruments Using a Weighted Median Estimator. Genet Epidemiol 40, 304–314 (2016). 10.1002/gepi.21965

104 Cheng, Q., Zhang, X., Chen, L. S. & Liu, J. Mendelian randomization accounting for complex correlated horizontal pleiotropy while elucidating shared genetic etiology. Nat Commun 13, 6490 (2022). 10.1038/s41467-022-34164-1

105 Morrison, J., Knoblauch, N., Marcus, J. H., Stephens, M. & He, X. Mendelian randomization accounting for correlated and uncorrelated pleiotropic effects using genome-wide summary statistics. Nat Genet 52, 740–747 (2020). 10.1038/s41588-020-0631-4

106 Lorincz-Comi, N., Yang, Y., Li, G. & Zhu, X. MRBEE: A bias-corrected multivariable Mendelian randomization method. HGG Adv 5, 100290 (2024). 10.1016/j.xhgg.2024.100290

107 Watanabe, K., Taskesen, E., van Bochoven, A. & Posthuma, D. Functional mapping and annotation of genetic associations with FUMA. Nat Commun 8, 1826 (2017). 10.1038/s41467-017-01261-5

108 Giambartolomei, C. et al. Bayesian test for colocalisation between pairs of genetic association studies using summary statistics. PLoS Genet 10, e1004383 (2014). 10.1371/journal.pgen.1004383

109 Ferkingstad, E. et al. Large-scale integration of the plasma proteome with genetics and disease. Nat Genet 53, 1712–1721 (2021). 10.1038/s41588-021-00978-w

110 Sun, B. B. et al. Plasma proteomic associations with genetics and health in the UK Biobank. Nature 622, 329–338 (2023). 10.1038/s41586-023-06592-6

111 Võsa, U. et al. Large-scale cis- and trans-eQTL analyses identify thousands of genetic loci and polygenic scores that regulate blood gene expression. Nat Genet 53, 1300–1310 (2021). 10.1038/s41588-021-00913-z

112 Soskic, B. et al. Immune disease risk variants regulate gene expression dynamics during CD4(+) T cell activation. Nat Genet 54, 817–826 (2022). 10.1038/s41588-022-01066-3

113 Yazar, S. et al. Single-cell eQTL mapping identifies cell type-specific genetic control of autoimmune disease. Science 376, eabf3041 (2022). 10.1126/science.abf3041

114 Zdrazil, B. et al. The ChEMBL Database in 2023: a drug discovery platform spanning multiple bioactivity data types and time periods. Nucleic Acids Res 52, D1180–d1192 (2024). 10.1093/nar/gkad1004

115 Chen, X., Ji, Z. L. & Chen, Y. Z. TTD: Therapeutic Target Database. Nucleic Acids Res 30, 412–415 (2002). 10.1093/nar/30.1.412

116 Finan, C. et al. The druggable genome and support for target identification and validation in drug development. Sci Transl Med 9 (2017). 10.1126/scitranslmed.aag1166

